# A nationwide joint spatial modelling of simultaneous epidemics of dengue, chikungunya, and Zika in Colombia

**DOI:** 10.1101/2024.10.25.24316124

**Authors:** Laís Picinini Freitas, Mabel Carabali, Alexandra M. Schmidt, Jorge Emilio Salazar Flórez, Brayan Ávila Monsalve, César García-Balaguera, Berta N. Restrepo, Gloria I. Jaramillo-Ramirez, Kate Zinszer

## Abstract

**Background:** Chikungunya, and Zika emerged in the 2010s in the Americas, causing simultaneous epidemics with dengue. However, little is known of these *Aedes*-borne diseases (ABDs) joint patterns and contributors at the population-level.

**Methods:** We applied a novel Poisson-multinomial spatial model to the registered cases of dengue (n=291,820), chikungunya (n=75,913), and Zika (n=72,031) by municipality in Colombia, 2014-2016. This model estimates the relative risk of total ABDs cases and associated factors, and, simultaneously, the odds of presence and contributors of each disease using dengue as a baseline category. This approach allows us to identify combined characteristics of ABDs, since they are transmitted by the same mosquitoes, while also identifying differences between them.

**Findings:** We found an increased ABDs risk in valleys and south of the Andes, the Caribbean coast, and borders, with temperature as the main contributor (Relative Risk 2.32, 95% Credible Interval, CrI, 2.05-2.64). Generally, dengue presence was the most probable among the ABDs, although that of Zika was greater on Caribbean islands. Chikungunya and Zika were more likely present than dengue in municipalities with less vegetation (Odds Ratio, OR, 0.75, 95%CrI 0.65-0.86, and 0.85, 95%CrI 0.74-0.99, respectively). Chikungunya tended to be present in more socially vulnerable areas than dengue (OR 1.20, 95%CrI 0.99-1.44) and Zika (OR 1.19, 95%CrI 0.95-1.48).

**Interpretation:** Important differences between the ABDs were identified and can help guide local and context-specific interventions, such as those aimed at preventing cases importation in border and tourism locations and reducing chikungunya burden in socially vulnerable regions.

## Introduction

Dengue affects tropical and subtropical regions, with an estimated 30-fold increase in incidence over the past 50 years and 6.5 million cases in 2023.^1,2^ Caused by four distinct viral serotypes and transmitted by *Aedes* mosquitoes, dengue control is difficult and epidemics occur cyclically.^3^ The last decades, dengue-endemic countries faced the emergence of other *Aedes*-borne diseases, notably chikungunya and Zika, leading to co-circulation and simultaneous epidemics, i.e. syndemics, a greater challenge to public health.^3–5^

Dengue, chikungunya, and Zika distribution at the population-level is associated with factors affecting the presence, density, and vector competence of *Aedes* mosquitoes, such as climate, vegetation, and sanitation.^6–8^ Warmer temperatures caused by climate change are also thought to drive dengue expansion to previously unaffected regions.^9–12^ Urban, densely populated, and socially vulnerable locations are considered at increased risk for *Aedes*-borne diseases, reflecting the mosquito’s adaptation to living close to humans.^8,13–15^

Because these arboviruses share the same vector and co-circulate, it is expected that their spatial distribution is correlated, advocating for a joint analysis. In Colombia, a study applied multivariate scan statistics to detect simultaneous clusters of dengue, chikungunya, and Zika.^16^ Although risk-areas for their joint presence were identified, clusters were also found for only one or two diseases at a time. We consider that certain population-level factors (e.g., humidity, sanitation, etc.) impact each arbovirus distribution differently.

We applied a joint multivariate spatial model^17^ to simultaneously study dengue, chikungunya, and Zika outbreaks in Colombia. This model provides the joint and individual measures of risk/probability of presence and measures of association with covariates. This represents a novel and meaningful approach, allowing the identification of distinct patterns and contributing factors for each disease while accounting for their similarities.

## Methods

This ecological study used national surveillance data of notified dengue, chikungunya, and Zika cases in Colombia grouped by municipality, from 2014 to 2016. Hereon, we use “*Aedes*-borne diseases” to encompass dengue, chikungunya, and Zika together.

We applied a Poisson-multinomial spatial model to estimate i) the relative risk of the total cases of *Aedes*-borne diseases, ii) the probability of presence of each disease, given the total, and ii) the association of covariates with the relative risk of *Aedes*-borne diseases and with the odds of presence of one disease in comparison to each other.

### Study site

Colombia is located in northern South America and its territory is divided into 1,121 municipalities, grouped into 33 departments (Supplementary Figure S1). Colombia borders five countries, has two coastlines, an archipelago in the Caribbean Sea, three mountain ranges forming the Colombian Andes, a savanna region east of the Andes, and an Amazon rainforest in the south.

### Data

Anonymized individual records of dengue (2014-2016), chikungunya (2014-2016), and Zika (2015-2016) cases were obtained from the Colombian National Public Health Surveillance System (SIVIGILA). Data correspond to individuals diagnosed with dengue, chikungunya, or Zika at a healthcare facility, using case definitions from Colombian National Health Institute protocols.^18–20^ Cases matching a suspected or confirmed case definition (Supplementary Text S1) and with symptom onset between January 3 2014 to October 1 2016 (epidemiological weeks, EWs, 01/2014 to 39/2016) were included and aggregated by municipality of residence.

Environmental data for the same period, including the Normalised Difference Vegetation Index (NDVI, which quantifies vegetation greenness, with values near +1 representing dense vegetation cover), mean temperature, relative humidity, and total precipitation by week and municipality, were obtained from data previously processed.^21,22^ We calculated the average values of these variables for each municipality (Supplementary Figure S2). Population estimates for 2014-2016 were obtained from Colombia’s National Administrative Department of Statistics (DANE) and used to calculate the population density per km^2^ (Supplementary Figure S3A). Socioeconomic data by municipality, including the percentage of people using rainwater for cooking, the percentage of homes with garbage collection, and the percentage of people with unsatisfied basic needs (UBN) were sourced from the national census of 2018 (DANE) (Supplementary Figure S3B-D). The UBN index captures socioeconomic vulnerabilities based on housing conditions, overcrowding, and access to basic sanitation, among others.^23^ Indexes for “real” (e.g., percentage of births attended by qualified personnel) and “potential” (e.g., number of healthcare beds and general practitioners) healthcare access for 2016-2017 were obtained from the Colombian National Health Observatory (ONS). These indexes range from 0 to 100, with 0 representing the worst healthcare access and 100 the best (Supplementary Figure S3E-F).^24^

### Statistical methods

We applied a Poisson-multinomial spatial model under the Bayesian framework, originally proposed by Schmidt et al.^17^ Let *D*_*i*_, *C*_*i*_ and *Z*_*i*_ denote the number of dengue, chikungunya, and Zika cases, respectively, in municipality *i* = 1, 2, …, n = 1121, during the study period. And let *total*_*i*_ = (*D*_*i*_ + *C*_*i*_ + *Z*_*i*_) be the total number of cases of the three *Aedes*-borne diseases. We assume that

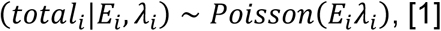

being *E*_*i*_ the number of expected cases considering a homogenous incidence rate across the municipalities, calculated as 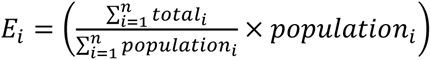, and *λ*_*i*_ the relative risk associated with the total number of cases of *Aedes*-borne diseases, modelled as

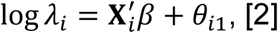

where 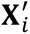 is a vector of covariates, including an intercept, *β* is a vector of coefficients, and *θ*_*i*1_ is a latent effect for municipality *i*.

Now let 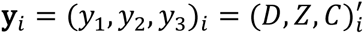 be a three-dimensional vector with the number of registered cases of each disease in municipality *i*. When total > 0:

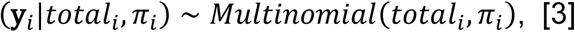

where *π*_*i*_ = (*π*_*i*1_, *π*_*i*2_, *π*_*i*3_)′ is the vector of probabilities of occurrence of dengue, chikungunya, or Zika in municipality *i*, with 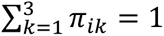. We assume that

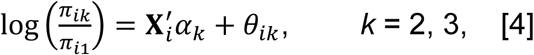

to model the conditional probability of chikungunya presence (*k*=2) given that it is chikungunya or dengue (*k*=1), and of Zika presence (*k*=3) given that it is Zika or dengue. Dengue was considered the baseline category due to endemicity before the introduction of chikungunya and Zika. The covariates’ coefficients *⍺*_*k*_ vary with *k* and exp (*⍺*_*kj*_) represents the odds ratio of chikungunya or Zika presence compared to dengue associated with one standard deviation increase in covariate *j*. The odds ratio associated with covariates and chikungunya presence compared to Zika is obtained from exp (*⍺*_2*j*_ − *⍺*_3*j*_), and vice-versa from exp (*⍺*_3*j*_ − *⍺*_2*j*_).

Note that the *θ*_*i*._ is present in equations [2] and [4]. Considering that the three diseases are transmitted by the same vectors, we assume that *θ*_*i*._ = (*θ*_*i*1_, *θ*_*i*2_, *θ*_*i*3_)^*T*^ ∼ *N*_3_[(1_3_)^*T*^*ϕ*_*i*_, Σ], i.e., within municipality *i*, the *θ*_*i*._’s share, *a priori*, a common spatial effect *ϕ*_*i*_, that follows a proper conditional autoregressive prior distribution.^25^

Models were fitted using the Stan platform^26^ in R (version 4.3.3), with the package rstan (version 2.32.6). Covariates were standardised. Convergence was checked by the R-hat statistic and by visually inspecting the chains. Figures were created using the packages ggplot2 (version 3.5.0) and colorspace (version 2.1-0) in R.

## Results

Between EWs 01/2014 and 39/2016, 291,820 dengue, 75,913 chikungunya and 72,031 Zika cases were registered in Colombia, totalling 439,769 cases of *Aedes*-borne diseases. The cumulative incidence at the national level was 629.8 cases per 100,000 inhabitants for dengue, 163.8 for chikungunya, 155.4 for Zika, and 949.1 for the three diseases. The incidence distribution by municipality is shown in Figure 1.

**Figure 1.**
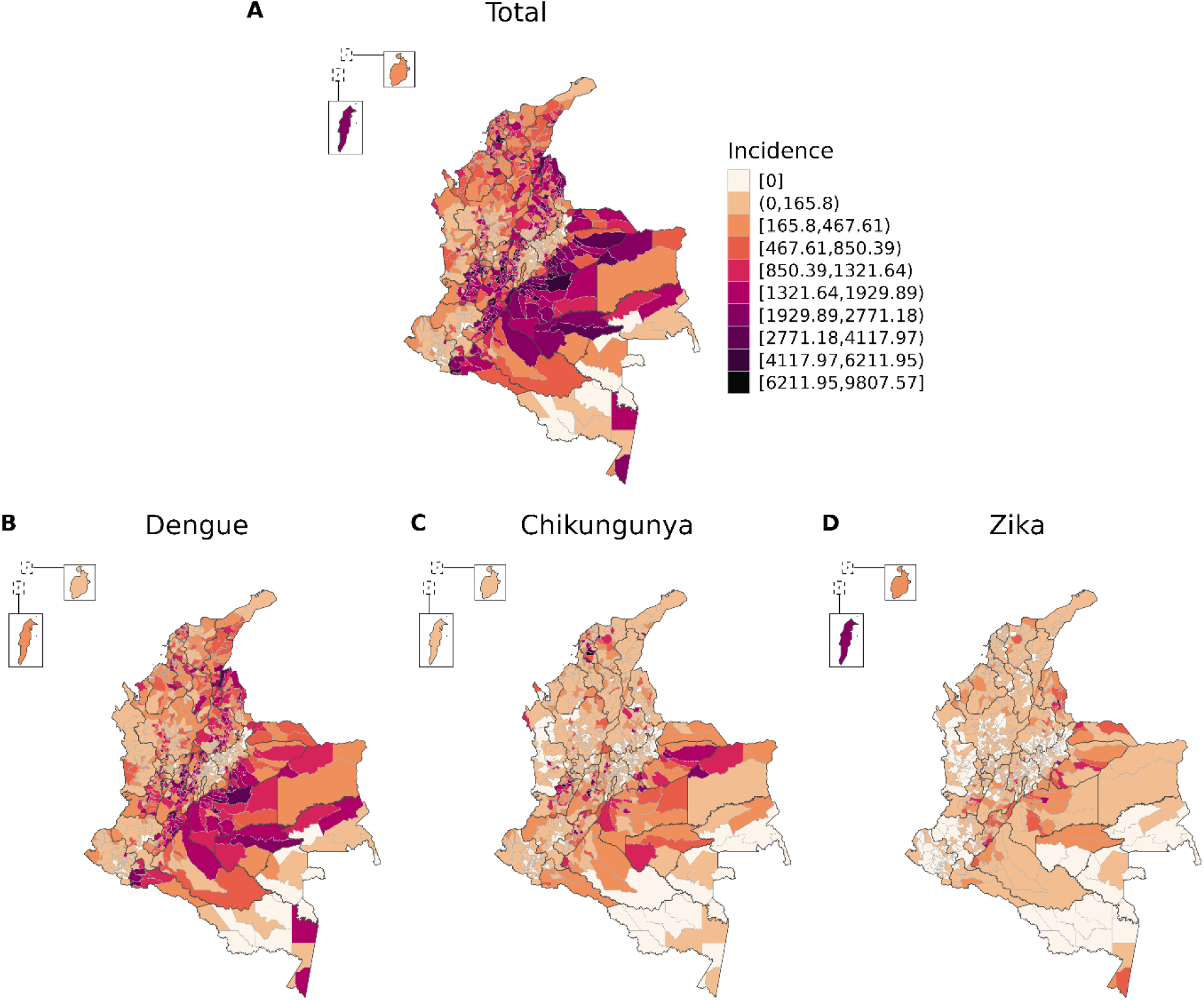
*Aedes*-borne diseases (dengue, chikungunya, and Zika) total incidence (per 100,000 inhabitants) and for each disease by municipality, Colombia, EW 01/2014 to 39/2016.

### Total relative risk and probability of presence of each disease

Figure 2A shows the relative risk for *Aedes*-borne diseases, i.e., for the total cases of the three diseases (dengue, chikungunya, and Zika) combined. Of the 1,121 municipalities, 357 (31.8%) had posterior mean relative risks >1. Higher risks were concentrated south of the Andean region (Meta, Casanare and Guaviare departments), in valleys in the Andean region (Tolima, Huila, Cundinamarca and inland Valle del Cauca), near the border with Venezuela (Santander and Norte de Santander) and Ecuador (Putumayo), in the Caribbean coast (Bolívar and San Andrés island), and in the southernmost municipality of the country, Leticia, that borders Brazil and Peru.

**Figure 2.**
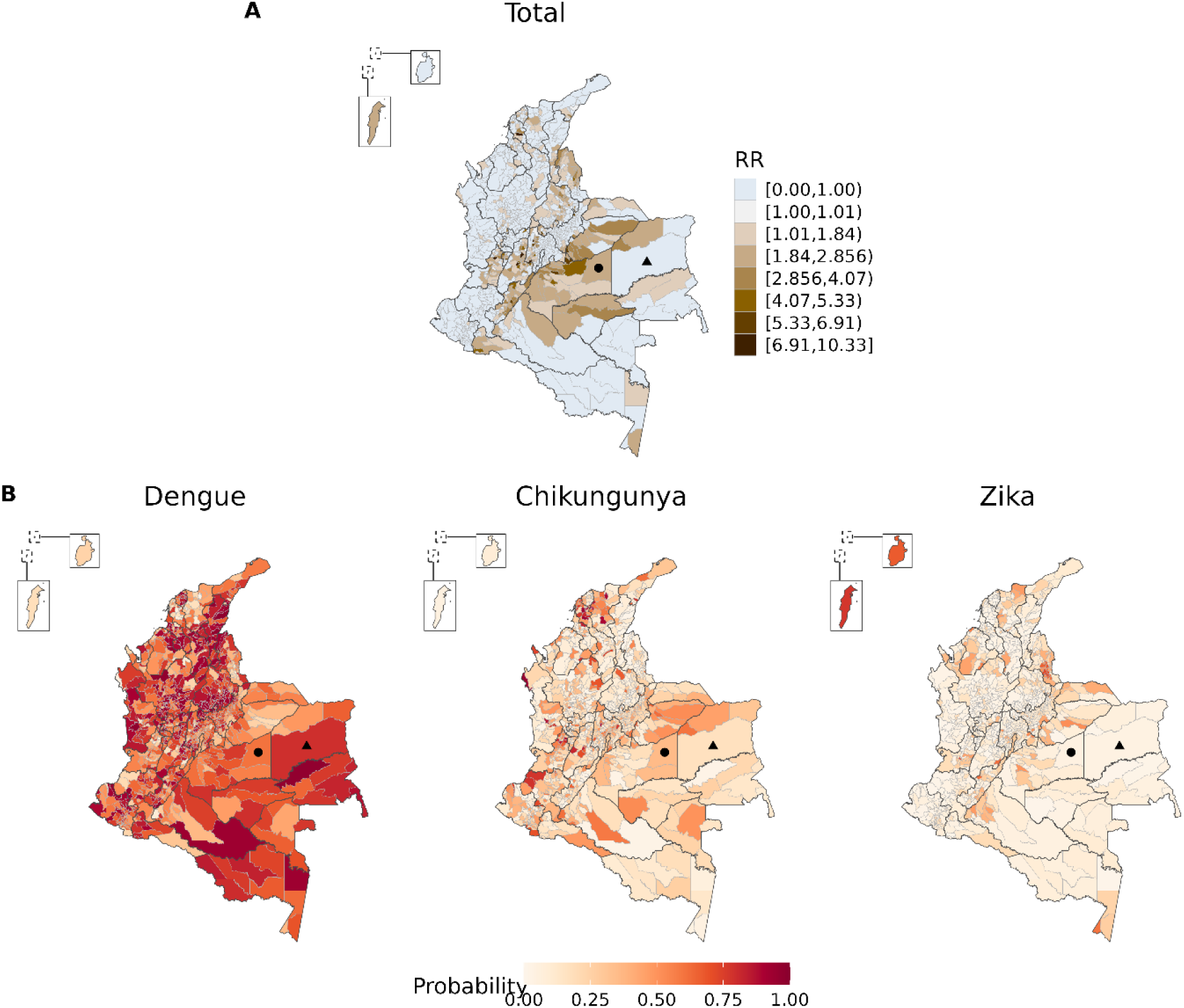
Posterior mean (A) of the relative risk (RR) for the total cases of *Aedes*-borne diseases (dengue, chikungunya, and Zika) and (B) of probabilities of presence of each disease, given the total, by municipality, Colombia, epidemiological week 01/2014 to 39/2016.

Figure 2B shows the conditional probability of presence of each disease. Generally, the probability of dengue presence was the highest among the arboviruses (Supplementary Figure S4). The probability of presence of one disease should be interpreted comparatively to the others and conditioned to the relative risk for the total *Aedes*-borne diseases. For example, from Figure 2A, Puerto Gaitán (solid circle) had an overall relative risk for *Aedes*-borne diseases >1 (above the expected for its population) and, comparatively from Figure 2B, the probability of dengue presence was higher than that of chikungunya and Zika. In contrast, Cumaribo (solid triangle) had an overall relative risk <1 (below the expected given its population). And, under low risk of *Aedes*-borne diseases, dengue was more likely present in Cumaribo than chikungunya and Zika.

### Latent effects

Figure 3A depicts the posterior mean of the shared spatial effect (*ϕ*_*i*_) and Figure 3B the disease-specific latent effects (*θ*_*i*._). The distribution of the spatial effect was similar to that of the posterior relative risk (Figure 2A). High values of *θ*_*i*2_ and/or *θ*_*i*3_ represent an increased probability of presence of chikungunya and/or Zika, respectively, compared to dengue, not explained by the covariates and the shared spatial effect.

**Figure 3.**
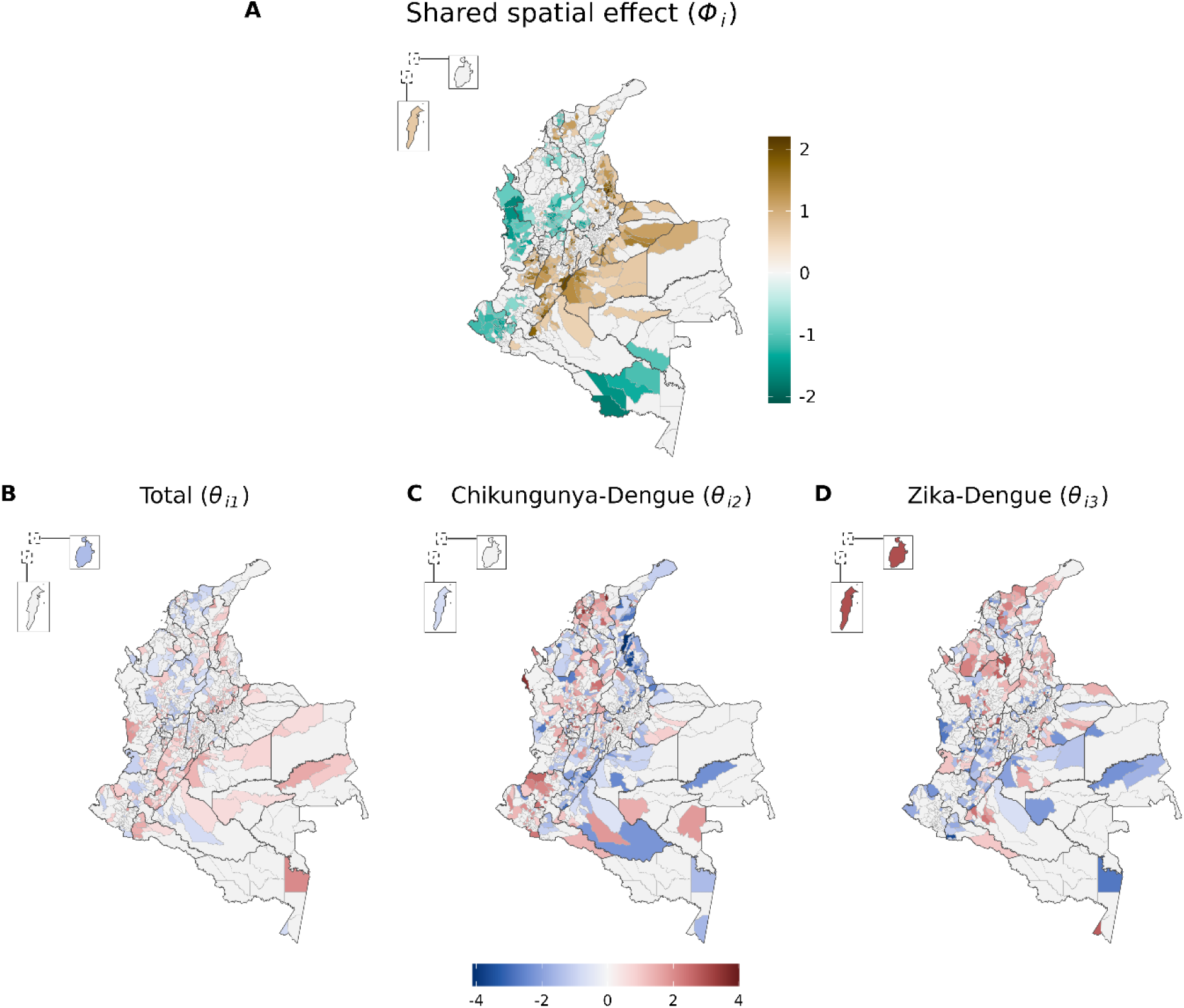
Posterior mean of the shared spatial effect for *Aedes*-borne diseases (dengue, chikungunya, and Zika) (A) and of the latent effects associated with the total cases (equation [2]) (B) and with the presence of chikungunya (C) or Zika (D) compared to dengue (equation [4]) by municipality in Colombia, epidemiological week 01/2014 to 39/2016. Municipalities for which the 95% credibility interval of the effect included 0 are shown in light grey.

### Association with covariates

The association with covariates is presented in Figure 4. Temperature showed the strongest association with the risk of *Aedes*-borne diseases (mean 2.32, 95% Credible Interval, CrI, 2.05-2.64). Warmer locations tended to have decreased odds of Zika presence compared to dengue (0.94, 95%CI 0.78-1.13) and chikungunya (0.90, 95%CI 0.74-1.09).

**Figure 4.**
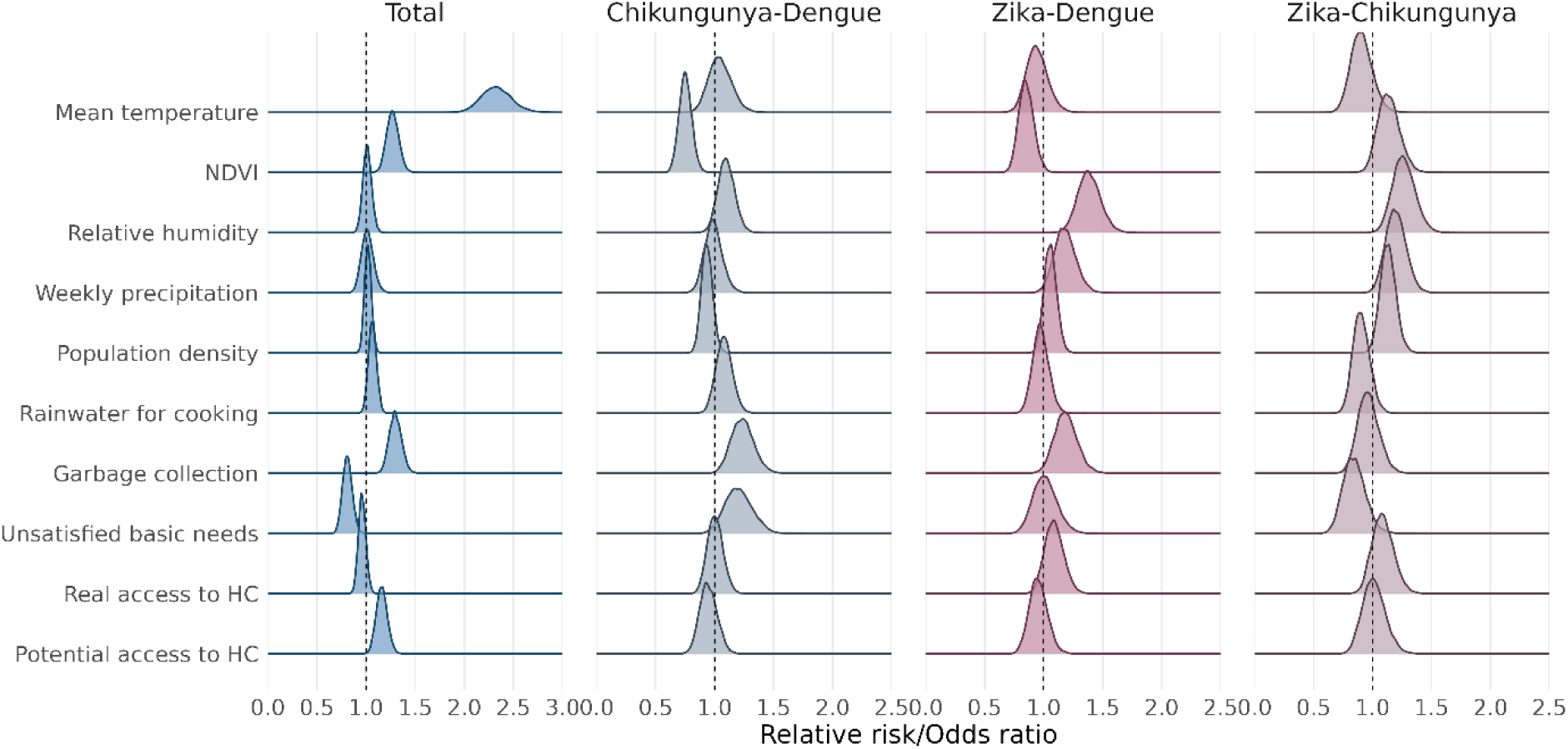
Posterior distribution of the relative risks associated with each of the covariates and the total number of cases of *Aedes*-borne diseases (dengue, chikungunya, and Zika), and of the odds ratios associated with each of the covariates and the odds of a municipality having chikungunya or Zika in comparison to dengue, and Zika in comparison to Chikungunya. NDVI = Normalized Difference Vegetation Index, HC = Healthcare.

The NDVI was positively associated with the total *Aedes*-borne diseases risk (1.27, 95%CrI 1.16-1.40), and inversely associated with the odds of chikungunya (0.75, 95%CrI 0.65-0.86) and Zika (0.85, 95%CrI 0.74-0.99) presence compared to dengue. Locations with higher NDVI had slightly increased odds of Zika compared to chikungunya (1.13, 95%CrI 0.97-1.33).

Humidity, precipitation, and population density were not associated with the *Aedes*-borne diseases risk. However, municipalities with higher humidity and/or weekly precipitation had increased odds of Zika presence compared to dengue (1.38, 95%CrI 1.21-1.58; and 1.18, 95%CrI 1.01-1.37, respectively) and chikungunya (1.26, 95%CrI 1.09-1.46; and 1.20, 95%CrI 1.04-1.37). More humid municipalities tended to have higher odds of chikungunya presence compared to dengue (1.10, 95%CrI 0.97-1.24). Densely populated municipalities had increased odds of having Zika compared to chikungunya (1.13, 95%CrI 1.01-1.27). These areas also tended to have increased odds of Zika presence (1.06, 95%CrI 0.96-1.17), and lower odds of chikungunya presence (0.93, 95%CrI 0.85-1.03) compared to dengue.

Municipalities with a higher percentage of people using rainwater for cooking tended to have a positive association with the total risk (1.07, 95%CrI 0.99-1.17), and higher odds of chikungunya presence compared to dengue (1.08, 95%CrI 0.95-1.22) and Zika (1.11, 95%CrI 0.95-1.29).

Municipalities with a higher percentage of homes with garbage collection presented a higher risk of total *Aedes*-borne diseases (1.30, 95%CrI 1.18-1.43), and higher odds of presence of chikungunya (1.23, 95%CrI 1.06-1.44) and Zika (1.19, 95%CrI 1.01-1.39) compared to dengue.

The UBN was inversely associated with the risk of *Aedes*-borne diseases (0.81, 95%CrI 0.72-0.91), and borderline associated with increased odds of chikungunya presence compared to dengue (1.20, 95%CrI 0.99-1.44) and Zika (1.19, 95%CrI 0.95-1.48).

Real access to healthcare was not associated with the risk of *Aedes*-borne diseases. Increased Zika presence probability was estimated in locations with better real access to healthcare compared to dengue (1.09, 95%CrI 0.94-1.26) and chikungunya (1.08, 95%CI 0.92-1.28). Finally, better potential healthcare access was positively associated with the total *Aedes*-borne diseases cases (1.16, 95%CrI 1.06-1.28), with no differences in the odds of presence between the diseases.

## Discussion

We used a novel syndemic approach to estimate the relative risk of *Aedes*-borne diseases and the conditional probability of presence of each disease. The overall spatial distribution of *Aedes*-borne diseases varied across Colombia. Temperature was the main contributor to arbovirus presence at the municipal level. To our knowledge, this is the first study that uses a joint spatial model for dengue, chikungunya, and Zika risk simultaneously for an entire country.

Increased risk of *Aedes*-borne diseases was estimated in regions historically burdened by dengue^27^ (in valleys and south of the Andes), tourist locations (in the Caribbean coast and islands), and near international borders. These regions also showed strong common spatial effects in agreement with the consideration of shared measured and unmeasured characteristics associated with the presence and distribution beyond the shared vector. Some of these regions were previously identified as high-risk areas for dengue using a Climate Risk Factor analysis^28^, as high-risk clusters for dengue, chikungunya, and Zika using Kulldorff’s scan statistics^16,29,30^, and with elevated burden for *Aedes*-borne diseases^31^.

The higher probability of dengue presence compared to chikungunya and Zika in most municipalities is likely due to dengue’s endemicity in Colombia, with the four serotypes in co-circulation.^32^ Conversely, chikungunya and Zika were emerging in the country. Importantly, Zika causes milder symptoms and is likely the most difficult *Aedes*-borne disease to detect by passive surveillance. Despite this, Zika was the one most likely present in the archipelago of San Andrés, Providencia y Santa Catalina, where thousands of tourists go annually and the disease was potentially introduced in Colombia.^16^

The temperature was identified as the main contributor to *Aedes*-borne diseases, which is supported by several other studies.^8,11,33–36^ Warmer temperatures accelerate both the extrinsic incubation period of arboviruses and the *Aedes* life cycle, boosting transmission and increasing mosquitoes population.^33^ Many Colombian municipalities located in mountains have been protected against arboviruses due to altitude, but rising temperatures from climate change may put them at risk.

We identified a positive association between NDVI and *Aedes*-borne diseases cases, likely driven by dengue, which was widespread in Colombia. Accordingly, more urbanised (i.e. lower NDVI) municipalities had a higher probability of chikungunya and/or Zika presence compared to dengue. These indicate the ruralisation of dengue in Colombia, similar to trends in other Latin American cities^10,37–39^, and the concentration of chikungunya and Zika first epidemics in more urbanised municipalities.

Despite dengue’s ruralisation, *Aedes*-borne diseases cases were still concentrated in large urban centres, where better socioeconomic indicators were often observed. This may explain the increased risk of *Aedes*-borne diseases in municipalities with a lower UBN and/or a higher percentage of homes with garbage collection. A similar positive association with garbage collection was found in Brazil.^40^ The authors discussed this indicator does not necessarily mean proper solid waste disposal, and disposal in open pits could increase mosquito breeding sites.

Interestingly, we found that, among the *Aedes*-borne diseases, chikungunya had a higher probability of presence in more socially vulnerable locations. Similar findings were described in a nationwide study in Brazil.^40^ Although this could result from disease spread from different entry sites, chikungunya may require more favourable transmission conditions, such as increased vector density and crowding, than dengue and Zika. This is supported by evidence that *Ae. aegypti* transmits chikungunya at lower rates than other arboviruses.^41,42^

Municipalities with better potential access to healthcare had more *Aedes*-borne diseases cases, likely reflecting better reporting and detection rates in these areas. A tendency of a higher probability of Zika presence compared to dengue and chikungunya was observed in locations with better real access to healthcare, which includes good prenatal and gynaecological services. Zika infection during pregnancy had been recently described to cause congenital malformations^43^, which probably increased awareness, and, in turn, the sensitivity of Zika detection among pregnant women of childbearing age. Also, nearly 90% of pregnant women attended at least four prenatal visits in Colombia in 2016,^44^ improving their access to the health system and infection detection compared to other groups.

Differences in the spatial distribution of disease presence probabilities could not be fully explained by the covariates. This advocates for local-specific interventions and preparing the healthcare system considering the epidemiological scenario and other local contexts. Strategies in tourist and border locations should aim at preventing the introduction of viruses, with active surveillance sensitive to detect asymptomatic and mild cases. Nevertheless, on the border with Ecuador and Venezuela, there is an important context of social vulnerability that such strategies must also alleviate.^45^ Collaboration with neighbouring countries in data sharing and strategy alignment is crucial, especially in twin cities like Letícia (Colombia) and Tabatinga (Brazil).

This study has limitations. Surveillance data are prone to underreporting, particularly in locations with limited access to healthcare.^46^ However, the Colombian surveillance system is robust, with mandatory reporting and national coverage. Also, given the quality of the *Aedes*-borne diseases surveillance system, its data is used for public health decision-making.^32^ To account for the potential differential reporting given access to healthcare, we included two indicators of healthcare access in our models. Surveillance data can be affected by misclassification of cases and syndemicity makes confirmation challenging. In our study, 29.0% of cases were laboratory-confirmed.

In conclusion, we present, in an unprecedented way, a nationwide robust and in-depth study of the syndemics of dengue, chikungunya, and Zika in Colombia using a multivariate model that allows comparative, yet simultaneous, analysis of disease distribution. This Poisson-multinomial spatial model can be applied in other *Aedes*-endemic countries, at other scales, and also for other diseases causing simultaneous outbreaks (e.g. influenza and COVID-19).

## Ethics approval

This study was approved by the Science and Health Research Ethics Committee (*Comité d’éthique de la recherche en sciences et en santé* - CERSES) of the University of Montreal, approval number CERSES-19-018-D.

## Data availability statement

The data used in this study are secondary data and are publicly available. Cases data are available at the Colombian National Public Health Surveillance System (*Sistema Nacional de Vigilancia en Salud Pública* - SIVIGILA) website (http://portalsivigila.ins.gov.co/). Environmental data was organised and made available by Siraj et al. (2019) (https://doi.org/10.5061/dryad.83nj1). Population and socioeconomic data are available at the National Administrative Department of Statistics of Colombia (*Departamento Administrativo Nacional de Estadística* - DANE) website (https://www.dane.gov.co/). A version of the healthcare access indexes can be found at the Colombian National Health Observatory (*Observatorio Nacional de Salud* - ONS) website (https://www.ins.gov.co/Direcciones/ONS/). Codes for the model are available at https://github.com/laispfreitas/joint_DZC_model.

## Author contributions

LPF conceptualised and designed the study; obtained, organised and analysed data; created figures; interpreted the results; wrote the manuscript. MC conceptualised and designed the study; interpreted the results; wrote the manuscript. AMS conceptualised and designed the study; analysed data; interpreted results; reviewed the manuscript. CGB, BNR and GIJR conceptualised the study; interpreted the results; reviewed the manuscript. KZ conceptualised and designed the study; interpreted the results; reviewed the manuscript.

## Acknowledgements

This work was supported by a grant from the Canadian Institutes of Health Research [grant number 428107]. Mabel Carabali holds a Fonds de recherche du Québec - Santé (FRQS) Chercheur Boursier Junior 1 [329874]. Alexandra M. Schmidt acknowledges financial support from the Natural Sciences and Engineering Research Council (NSERC) of Canada (Discovery Grants) [RGPIN-2024-04312]. Kate Zinszer holds a FRQS Chercheur Boursier Junior 2 [310728]. The funders had no role in study design, data collection and analysis, decision to publish, or preparation of the manuscript.

The authors would like to thank the *Instituto Nacional de Salud* of Colombia for making the diseases’ surveillance data publicly available, and the *Observatorio Nacional de Salud*, in particular Gina Alexandra Vargas Sandoval, Carlos Andres Castañeda Orjuela and Karol Patricia Cotes Cantillo, for sharing the healthcare access indexes data and for all their help with understanding and interpreting them.

## Declaration of interests

None.

## Notes

### Competing Interest Statement

The authors have declared no competing interest.

### Author Declarations

This study was approved by the Science and Health Research Ethics Committee (Comité d′é thique de la recherche en sciences et en santé - CERSES) of the University of Montreal, approval number CERSES-19-018-D.

