## Supplementary Material for "A nationwide joint spatial modelling of simultaneous epidemics of dengue, chikungunya, and Zika in Colombia"

**Supplementary Figure S1.** Colombia localisation and geographical departments.

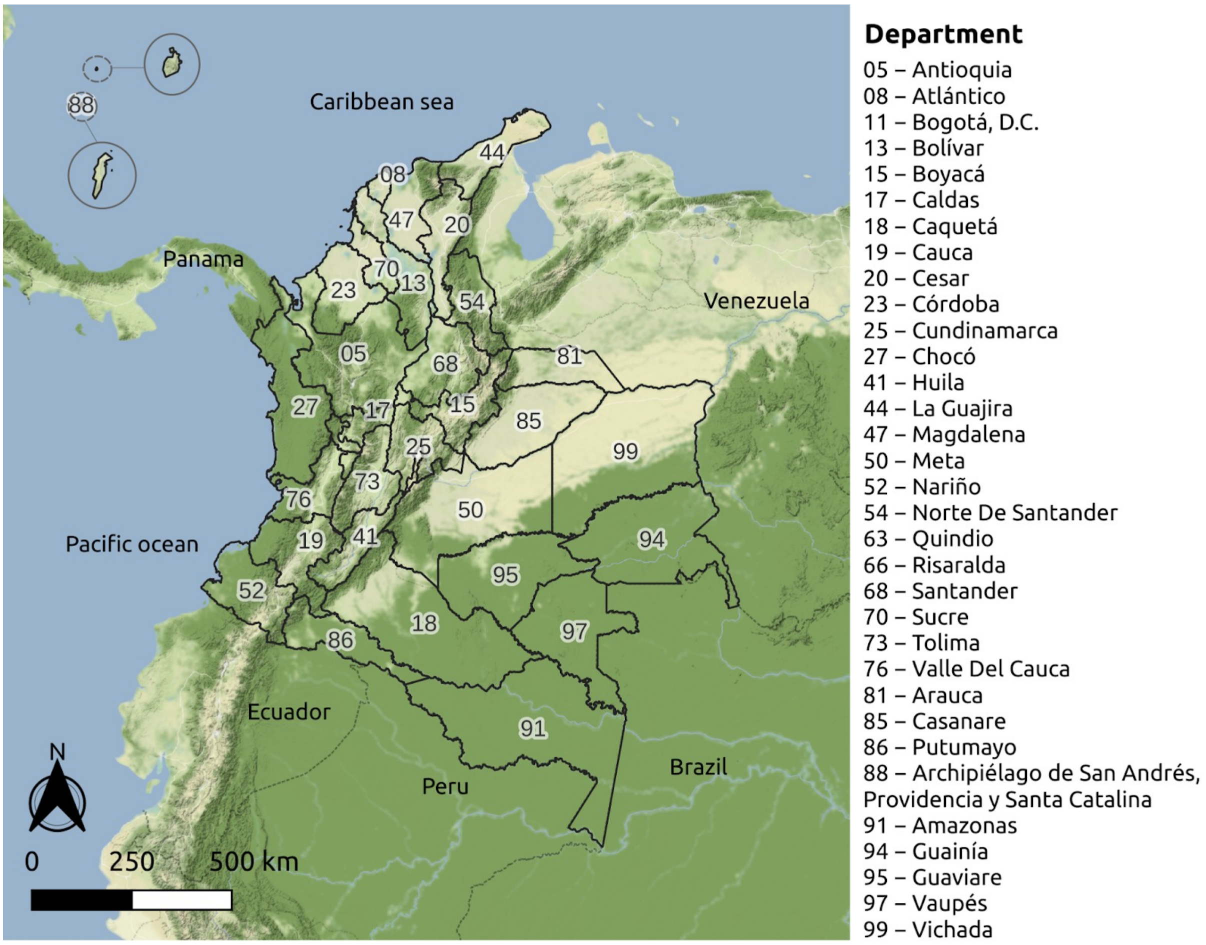


Map created using QGIS (version 3.22) (QGIS.org, 2021. QGIS Geographic Information System. QGIS Association. http://www.qgis.org). Sources: Colombian National Administrative Department of Statistics - DANE - geoportal. Background map tiles by Stamen Design, under CC BY 3.0. Data by OpenStreetMap, under ODbL.

**Supplementary Figure S2.** Environmental climate covariates by municipality. Mean values for (A) mean temperature, (B) the Normalised Difference Vegetation Index (NDVI), (C) relative humidity, (D) weekly precipitation, epidemiological week 01/2014 to 39/2016, Colombia.

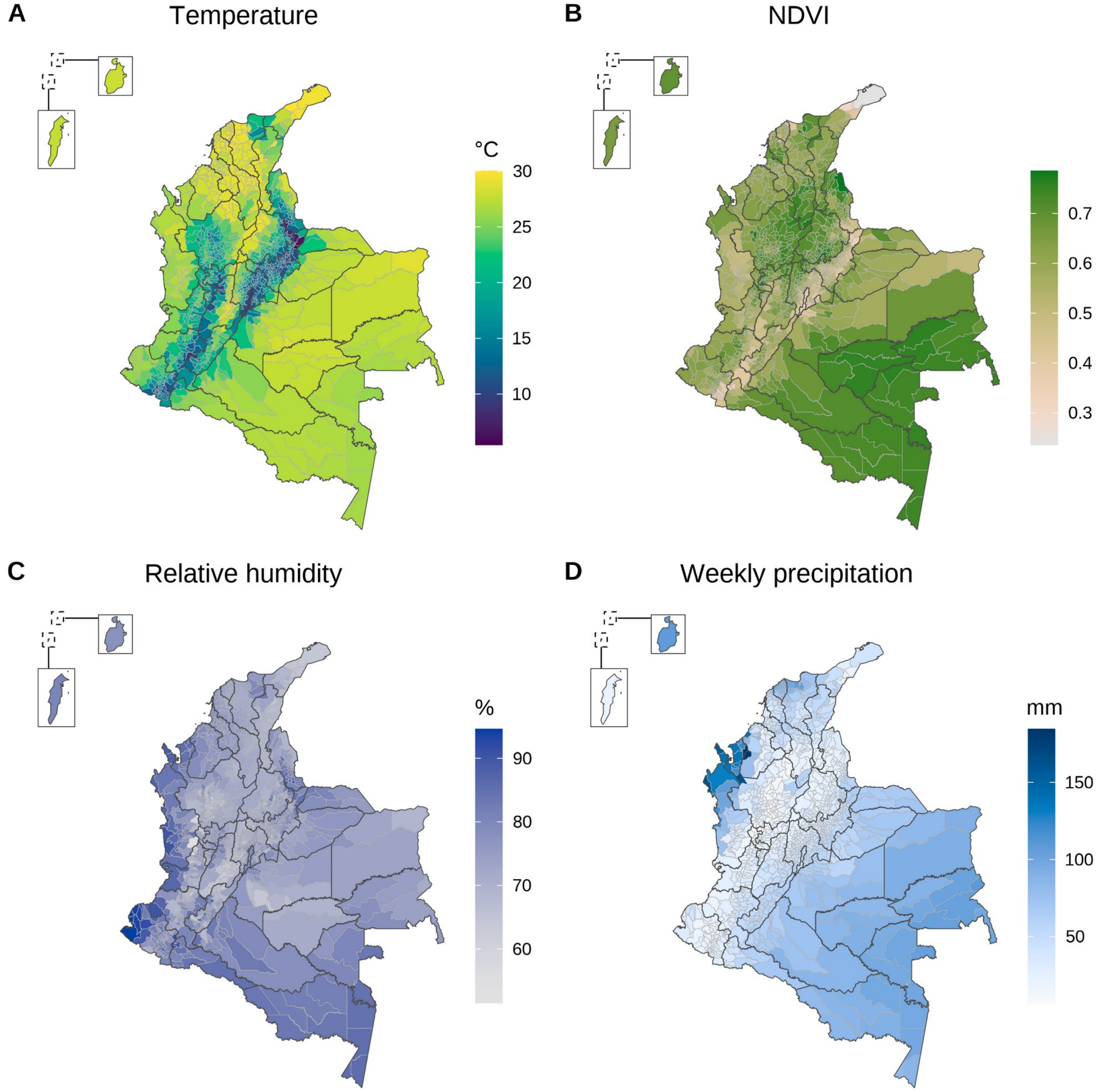


Maps created using R (version 4.3.3, https://www.r-project.org/). Data sources: Colombian National Administrative Department of Statistics - DANE; Siraj et al. (2019).

**Supplementary Figure S3.** Sociodemographic covariates by municipality: (A) mean population density (2014-2016), (B) percentage of people using rainwater for cooking (2018), (C) percentage of homes with garbage collection (2018), (D) percentage of population with unsatisfied basic needs (2018), (E) potential access to healthcare index (2016-2017), and (F) real access to healthcare index (2016-2017), Colombia.

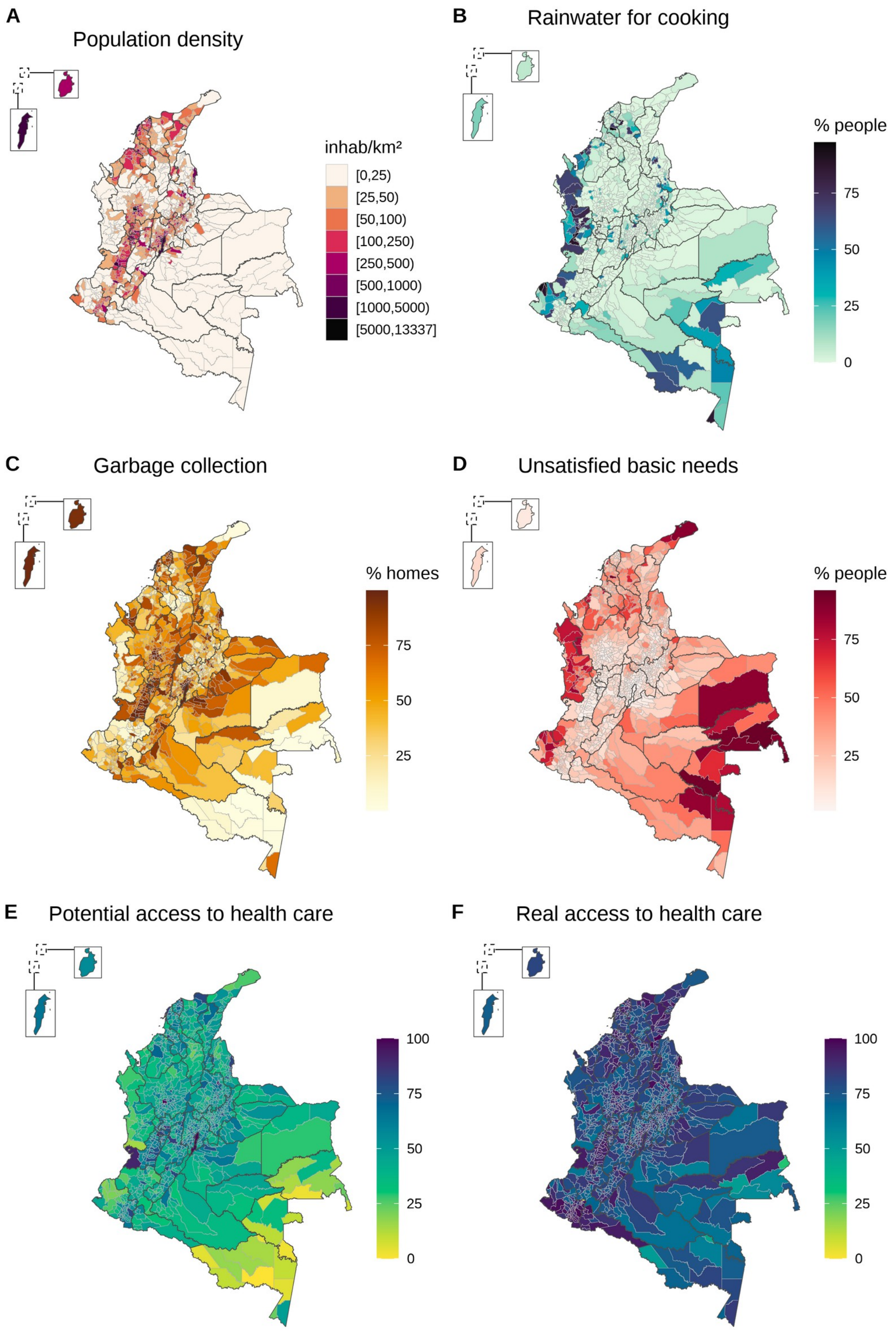


Maps created using R (version 4.3.3, https://www.r-project.org/). Data sources: Colombian National Administrative Department of Statistics - DANE; Colombian National Health Observatory - ONS.

**Supplementary Figure S4.** Map depicting the *Aedes*-borne disease (dengue, chikungunya, or Zika) with the highest posterior mean probability of presence in relation to the others by municipality, Colombia, epidemiological week 01/2014 to 39/2016.

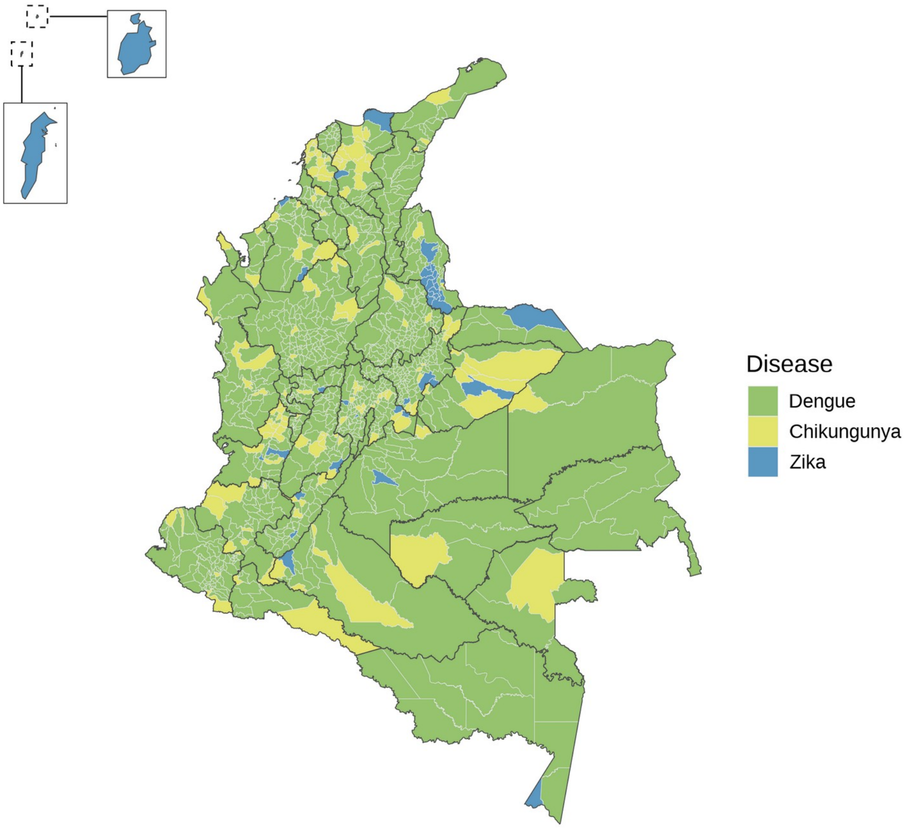


Map created using R (version 4.3.3, https://www.r-project.org/).

### Supplementary Text S1

#### Dengue case definition

The following criteria describe the case definition for dengue cases adopted by the Colombian National Institute of Health (INS).[1] The INS uses the word "probable" for dengue cases definition, and "suspected" for chikungunya and Zika. In this context, both terms have the same meaning. Therefore, to maintain consistency in the definition of the three diseases, we translated it to "suspected".

- Suspected case: Patient from an endemic area who meets the definition of dengue with or without warning signs.

Dengue without warning signs: Acute febrile illness of 2 to 7 days of evolution in which two or more of the following manifestations are observed: headache, retro-ocular pain, myalgia, arthralgia, skin eruption, rash or leukopenia.

Dengue with warning signs: Patient who meets the above definition and also presents any of the following warning signs: Intense and continuous abdominal pain or tenderness, persistent vomiting, diarrhoea, fluid accumulation (ascites, pleural effusion, pericardial effusion), mucosal bleeding, lethargy or irritability (mainly in children), postural hypotension, painful hepatomegaly >2 cm, drop in temperature, abrupt drop in platelets (<100,000) associated with hemoconcentration.

- Suspected case of severe dengue: Any case of dengue that meets any of the following serious manifestations of dengue: severe extravasation of plasma leading to dengue shock syndrome or fluid accumulation with respiratory distress; severe bleeding with hemodynamic compromise; clinical or paraclinical signs of severe organ damage such as liver damage, central nervous system damage, heart or other organ involvement.

- Case confirmed by epidemiological criteria: Suspected case residing within a perimeter of 200 metres (approximately two blocks) of another laboratory-confirmed case within 21 days (3 weeks) before or after laboratory diagnosis.

- Case confirmed by laboratory: Suspected case of dengue, severe dengue, or death due to dengue confirmed by any of the laboratory criteria for the diagnosis of dengue. PCR or viral isolation in patients with less than 5 days of onset of fever or IgM Dengue ELISA test in patients with 5 or more days of onset of fever (rapid tests are not accepted).

- Discarded case: A suspected case in which a laboratory test was carried out within the time established for the detection of the viral agent, and showed negative results for dengue virus, and another etiological diagnosis was established.

**Chikungunya case definition**

The following criteria describe the case definition for chikungunya cases adopted by the INS.[2]

- Suspected case: Patient who resides or has visited 8 to 15 days before the onset of symptoms, a municipality located between 0 and 2,200 metres above sea level, where no cases of chikungunya have been confirmed by laboratory and who has a fever greater than 38°C, severe arthralgia or acute-onset arthritis, erythema multiforme, or symptoms not explained by other medical conditions. Risk group patient who comes from areas located between 0 and 2,200 metres above sea level (regardless of whether or not they have confirmed viral circulation), 8 to 15 days before the onset of symptoms, who has a fever greater than 38°C, severe arthralgia or acute-onset arthritis and erythema multiforme or symptoms that are not explained by other medical conditions and from whom the sample is collected.

- Case confirmed by clinical epidemiological criteria: Patient with fever greater than 38°C, severe arthralgia or arthritis of acute onset, erythema multiforme or symptoms that are not explained by other medical conditions, who resides in or has visited a municipality where there is evidence of the circulation of the chikungunya virus, or is located in a municipality with a radius of 30 kilometres to municipalities with viral circulation.

- Case confirmed by laboratory: Probable case with any of the following virus-specific laboratory tests with a positive result (viral isolation, RT-PCR, IgM), or a four-fold increase in the titer of specific IgG antibodies to chikungunya virus in paired samples with a difference of 15 days between taking these.

- Discarded case: A suspected case in which a laboratory test was carried out and showed negative results for chikungunya virus, and another etiological diagnosis was established.

**Zika case definition**

The following criteria describe the case definition for Zika cases adopted by the INS.[3]

- Suspected case: Patient living in municipalities without confirmed Zika transmission presenting with a rash and one or more of the following symptoms not explained by other medical conditions: fever not greater than 38.5°C, nonpurulent conjunctivitis or conjunctival hyperemia, arthralgia, myalgia, headache, or malaise. Additionally, one of the following conditions: i) Person who visited, two weeks before the onset of symptoms, in countries or municipalities located between 0 and 2,200 m above sea level, with or without confirmed indigenous circulation of the Zika virus; ii) Person who had sexual contact without barrier protection two weeks before the onset of symptoms with a person who in the eight weeks prior to sexual contact visited areas with confirmed Zika transmission and/or areas with the presence of *Aedes* mosquitoes.

- Case confirmed by clinical epidemiological criteria: Patient who had been in countries or municipalities located between 0 and 2,200 metres above sea level with confirmed autochthonous circulation of the Zika virus two weeks before the onset of symptoms and who presented a rash and one or more of the following symptoms that were not explained by other medical conditions: fever not higher than 38.5°C, non-purulent conjunctivitis or conjunctival hyperemia, arthralgia, myalgia, headache or general malaise.

- Case confirmed by laboratory: Case that met the definition for probable or confirmed by clinical epidemiological case and that presented a positive result for Zika virus by RT-PCR (or immunohistochemistry in histopathological analysis) performed at the National Reference Laboratory of the NIH or collaborating centres designated by the NIH.

- Discarded case: A suspected case in which a laboratory test was carried out within the time established for the detection of the viral agent, and showed negative results for Zika virus, or another etiological diagnosis was established.
